# Prevalence of IgG antibodies against the severe acute respiratory syndrome coronavirus-2 among healthcare workers in Tennessee during May and June, 2020

**DOI:** 10.1101/2020.11.12.20230912

**Authors:** Peter F. Rebeiro, Kara J. Levinson, Lindsay Jolly, Elizabeth Kassens, George J. Dizikes, Richard S. Steece, David C. Metzger, Matthew Loos, Ron Buchheit, Lisa D. Duncan, Lori A. Rolando, Jonathan Schmitz, Heather A. Hart, David M. Aronoff, on behalf of the Tennessee COVID-19 Serology Study Team

## Abstract

SARS-CoV-2 seroprevalence was low (<1%) in this large population of healthcare workers (HCWs) across the state of Tennessee (n=11,787) in May-June 2020. Among those with PCR results, 81.5% of PCR and antibody test results were concordant. SARS-CoV-2 seroprevalence was higher among HCWs working in high-community-transmission regions and among younger workers.

**Importance:** These results may be seen as a baseline assessment of SARS-CoV-2 seroprevalence among HCWs in the American South during a period of growth, but not yet saturation, of infections among susceptible populations. In fact, this period of May-June 2020 was marked by the extension of renewed and sustained community-wide transmission after mandatory quarantine periods expired in several more populous regions of Tennessee. Where community transmission remains low, HCWs may still be able to effectively mitigate SARS-CoV-2 transmission, preserving resources for populations at high risk of severe disease, and these sorts of data help highlight such strategies.

## Background

Infection by the severe acute respiratory syndrome coronavirus-2 (SARS-CoV-2) causes coronavirus infectious disease from 2019 (COVID-19) (1) and is associated with the development of a humoral immune response, measurable by the presence of circulating immunoglobulins (Ig) G and M (2). Although the extent to which serological status against SARS-CoV-2 correlates with immunity against future infection remains uncertain (3), there is utility in using antibody testing to estimate the prevalence of infection within select populations, including healthcare workers (HCWs) (4).

The first laboratory confirmed case of COVID-19 in Tennessee was reported on March 4^th^, 2020. By mid-June there had been more than 30,000 cases diagnosed statewide (5). During this period, the Tennessee Department of Health partnered with healthcare systems belonging to the Tennessee Public and Teaching Hospital Association to estimate serological status of employees using a chemiluminescent immunoassay for IgG against SARS-CoV-2 (6). “Frontline” HCWs and employees previously diagnosed with COVID-19 were targeted.

## Methods

### Study population

HCWs ≥18 years of age were recruited and enrolled in the study between mid-May, 2020 and mid-June, 2020 at Erlanger Health System (Chattanooga, TN), Ballad Health (northeast TN), University of Tennessee Medical Center (UTMC, Knoxville, TN), Vanderbilt Wilson County Hospital (Lebanon, TN) and Vanderbilt University Medical Center (VUMC, Nashville, TN) sites (the latter two analyzed as VUMC). Ballad Health sites encouraged testing of all HCWs with direct patient contact while other sites recruited HCWs through convenience sampling among those in COVID-19 units, emergency departments, outpatient COVID-19 testing sites, and other service areas with high SARS-CoV-2 exposure probability.

### Sample collection & laboratory methods

Whole blood specimens were collected and an immunoassay for qualitative detection of anti-SARS-CoV-2 nucleocapsid protein-directed IgG in human serum and plasma was performed on these samples using the Abbott Architect i2000 with chemiluminescent microparticle immunoassay technology (7).

### Outcomes

The primary outcome was positive IgG result, reported within 7 days of sample collection. Among a sub-sample, PCR test results were self-reported on laboratory requisition forms or extracted from a surveillance database within the Tennessee Communicable and Environmental Diseases and Emergency Preparedness program.

### Covariates

Dates of sample collection and result reporting were used to assess trends in test volume. Outcomes and differences in age at the date of sample collection and sex assigned at birth (male or female) were also summarized by study enrollment site (as available).

### Statistical analysis

Non-parametric tests were used to describe contrasts in categorical and continuous variables across categorical variables such as serostatus, study site, and sex. For differences in proportions, the Pearson χ^2^ test was used; for differences in distributions, Wilcoxon rank-sum and Kruskal-Wallis tests were used. Age distributions were illustrated by violin plots. Temporal trends in testing were illustrated by kernel density plots using an epanechnikov kernel. Analyses were conducted using Stata v.15.1 (StataCorp, College Station, TX).

### Ethical clearance

A study exemption determination was obtained from the Tennessee Department of Health Institutional Review Board.

## Results

Among 11,787 individuals participating, most had study-related IgG tests conducted in mid-May to mid-June 2020 (**Figure 1a**); 116 (0.98%) were seropositive for SARS-CoV-2 by IgG, though the proportion IgG+ was significantly different across facilities, with higher proportions IgG+ at UTMC and VUMC (1.3% and 2.6%, respectively, vs. ≤1% everywhere else, χ^2^ p-value<0.001; **Figure 1b**).

**Figure 1.**
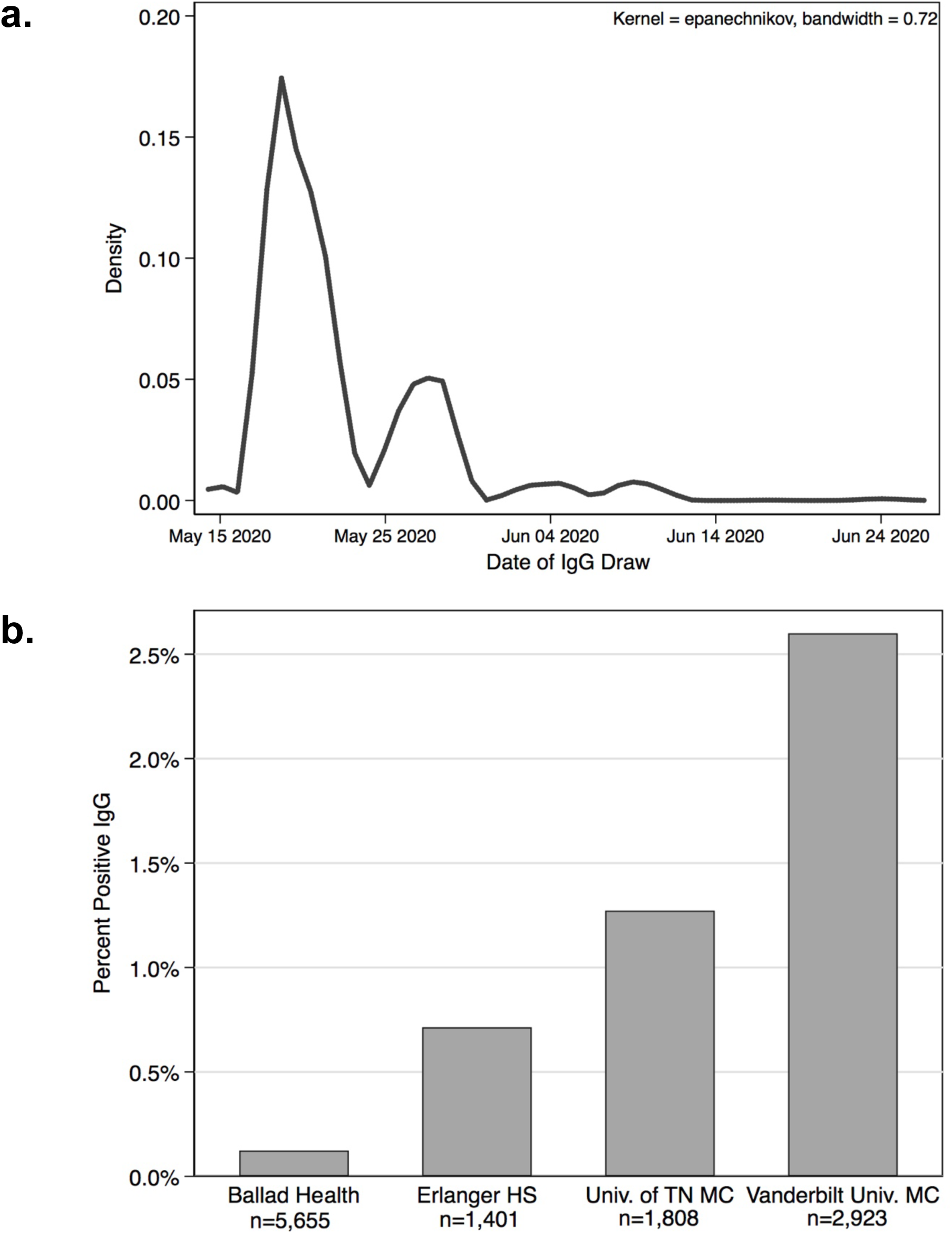
Distribution of collection dates for study-related SARS-CoV-2 serology (IgG) (**a**), and seropositivity by testing site (**b**), among 11,787 Tennessee healthcare workers, from May through June 2020

Median ages among those testing IgG- and IgG+ were 40.3 (interquartile range [IQR]: 30.9, 52.4) and 32.4 (IQR: 26.5, 43.4) years, respectively (Kruskal-Wallis p-value<0.001; **Figure 2a**).

**Figure 2.**
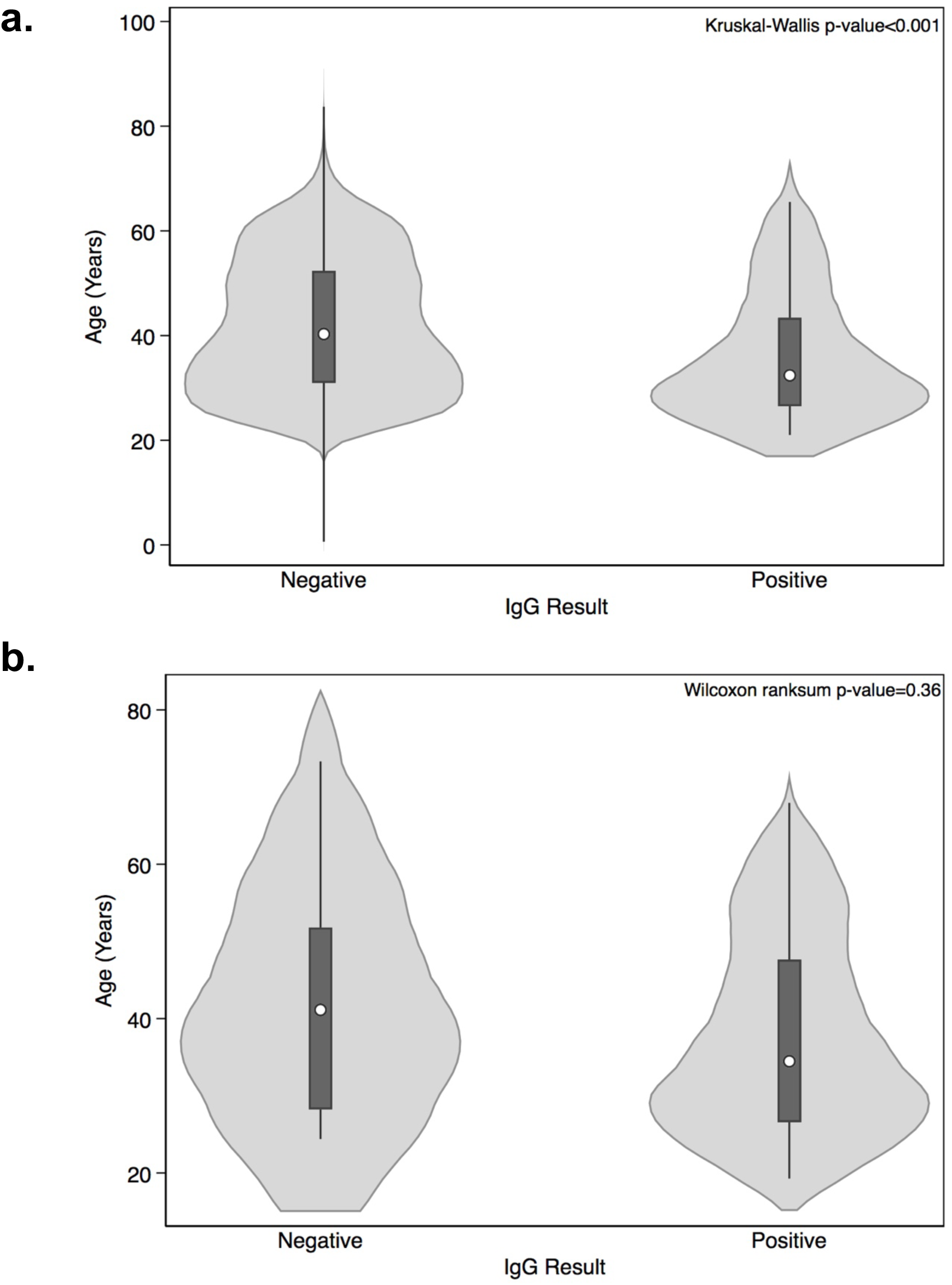
Violin plot describing the age distribution of participants by SARS-CoV-2 serostatus, among all 11,787 Tennessee healthcare workers participating (**a**.) and among 146 Tennessee healthcare workers who had both PCR and IgG testing available (**b**.), from May through June 2020

Among 6,108 individuals with sex assigned at birth available, 77% were female and 23% male. There were no significant differences by sex, with 22.8% and 22.0% male among IgG- and IgG+ participants, respectively (χ^2^ p-value=0.86).

Among 146 individuals who also had self-reported PCR testing for SARS-CoV-2, the lapses between PCR and IgG test dates were available for 54 individuals. Among participants with PCR test histories available, their first SARS-CoV-2 diagnostic tests (whether PCR or IgG) were collected from early March 2020 through June 2020, with most pre-study PCR testing done in mid-March and most IgG testing done in May; the distribution of lapses showed most PCR testing preceded study-related IgG testing by approximately 2 months (median lapse of 55 days).

Ages at sample collection were available for 143 individuals (97.9%) who had PCR test history. In this group, median age was 41.1 (IQR: 28.2, 51.8) years among those who were IgG- and 34.5 (IQR: 26.6, 47.6) years among those who were IgG+ (**Figure 2b**).

Among those ever PCR+, the proportion IgG+ was largely in agreement (percent agreement=81.5%), though they were not completely concordant: 3 of 54 individuals (5.6%) with test dates for both were IgG-even though they reported being PCR+ on ≥1 test, and 7 of 54 (13.0%) were IgG+ though they had reported only having PCR-tests (**Supplemental Figure 1**).

## Discussion

In a broad sero-survey of nearly 12,000 HCWs in systems across Tennessee during May-June 2020, the overall seropositivity for SARS-CoV-2 was low, approaching 1%. There was significant variation in seropositivity between healthcare systems, with higher seropositivity among workers in larger urban centers in middle and eastern Tennessee (Nashville and Knoxville). Seropositive persons were significantly younger than seronegative individuals, though there were no sex differences.

The depth and breadth of the regional COVID-19 epidemic in the American South had not yet made itself felt during the period of this particular sero-survey (8, 9). A smaller seroprevalence study (including 249 HCWs) from Nashville, conducted in April, 2020, found a higher seroprevalence (7.6%) but focused on high-risk hospital settings and detected any immunoglobulin isotype, including IgM, IgG, and IgA (10).

These results may be seen as a baseline assessment of SARS-CoV-2 seroprevalence among HCWs in the American South during a period of growth, but not yet saturation, of infections among susceptible populations. In fact, this period of May was marked by the extension of renewed and sustained community-wide transmission after mandatory quarantine periods expired in several more populous regions of Tennessee (5). Though the statewide average of newly detected infections was 5.49 per 100,000, Davidson county (middle Tennessee, home to VUMC) saw 10-20 new cases per 100,000 population; by contrast, Knox (northeast Tennessee, home to Ballad Health and UTMC) and Hamilton (southeast Tennessee, home to Erlanger Health System) counties saw only 1-5 new cases per 100,000 population in the same period. As may be expected, these pronounced regional differences in community transmission were reflected in differing levels of seropositivity among the corresponding HCW populations.

There were limitations in this study. First, differences in sampling schemes across health systems, and the voluntary participation of particular health systems, may render this study population not representative of the entire population of HCWs in the state of Tennessee, limiting generalizability. There also may have been outcome misclassification present, as no test, has perfect diagnostic sensitivity and specificity (7). Finally, this study did not have information on social network structures or other characteristics to identify high-risk groups for SARS-CoV-2 exposure and transmission among HCWs.

Seroprevalence patterns among HCWs, inherently at higher risk of exposure, might change as local and regional epidemics evolve and might be impacted by disease activity in the community, exposure mitigation efforts, personal protective equipment supplies, and public health policies (quarantines, mask mandates, etc.). The nature of SARS-CoV-2 transmission merits continued monitoring in this population in both the long and short-term.

## Data Availability

Data may not be made publicly available, as it contains identifiable and clinical data from participating healthcare workers otherwise collected for surveillance purposes.

## Acknowledgments & Conflicts of Interest

PFR received grant funding from NIH/NIAID (K01-AI131895). All authors report no other conflicts beyond standard NIH grant funding (money paid to their institutions). This work has not been presented at any meetings or in any other publications.

## Tennessee COVID-19 Serology Study Team

Katie Burger (Vanderbilt University Medical Center), Asimwe Baggett (TN Dept of Health, Div. of Laboratory Services), Alice C. Coogan (Vanderbilt University Medical Center), Assiatou Dialla (TN Dept of Health, Div. of Laboratory Services), Renee Grayson-Eubanks (Erlanger Health System), Chad Fitzgerald (Vanderbilt University Medical Center), Andrew Haag (Vanderbilt University Medical Center), Clisby Hall (Vanderbilt University Medical Center), April Kapu (Vanderbilt University Medical Center), Vicki Lambert (TN Dept of Health, Div. of Laboratory Services), Shelby Lowrie (TN Dept of Health, Div. of Laboratory Services), Bryan Mason, MS (TN Dept of Health, Div. of Laboratory Services), Stacy Prater (Erlanger Health System), Catherine Qian (Vanderbilt University Medical Center), Adam Seegmiller (Vanderbilt University Medical Center) Amanda Uhls (TN Dept of Health, Div. of Laboratory Services), Lily Vaden (TN Dept of Health, Div. of Laboratory Services).

## Figure Legends

**Supplemental Figure 1**. Venn diagram depicting the overlap in IgG+ and self-reported PCR+ SARS-CoV-2 test results (81.5% agreement), among 146 Tennessee healthcare workers who had both PCR and IgG testing available, from May through June 2020

