## Supplementary figures and images for "Prevalence of IgG antibodies against the severe acute respiratory syndrome coronavirus-2 among healthcare workers in Tennessee during May and June, 2020"

### Supplemental Figure 1

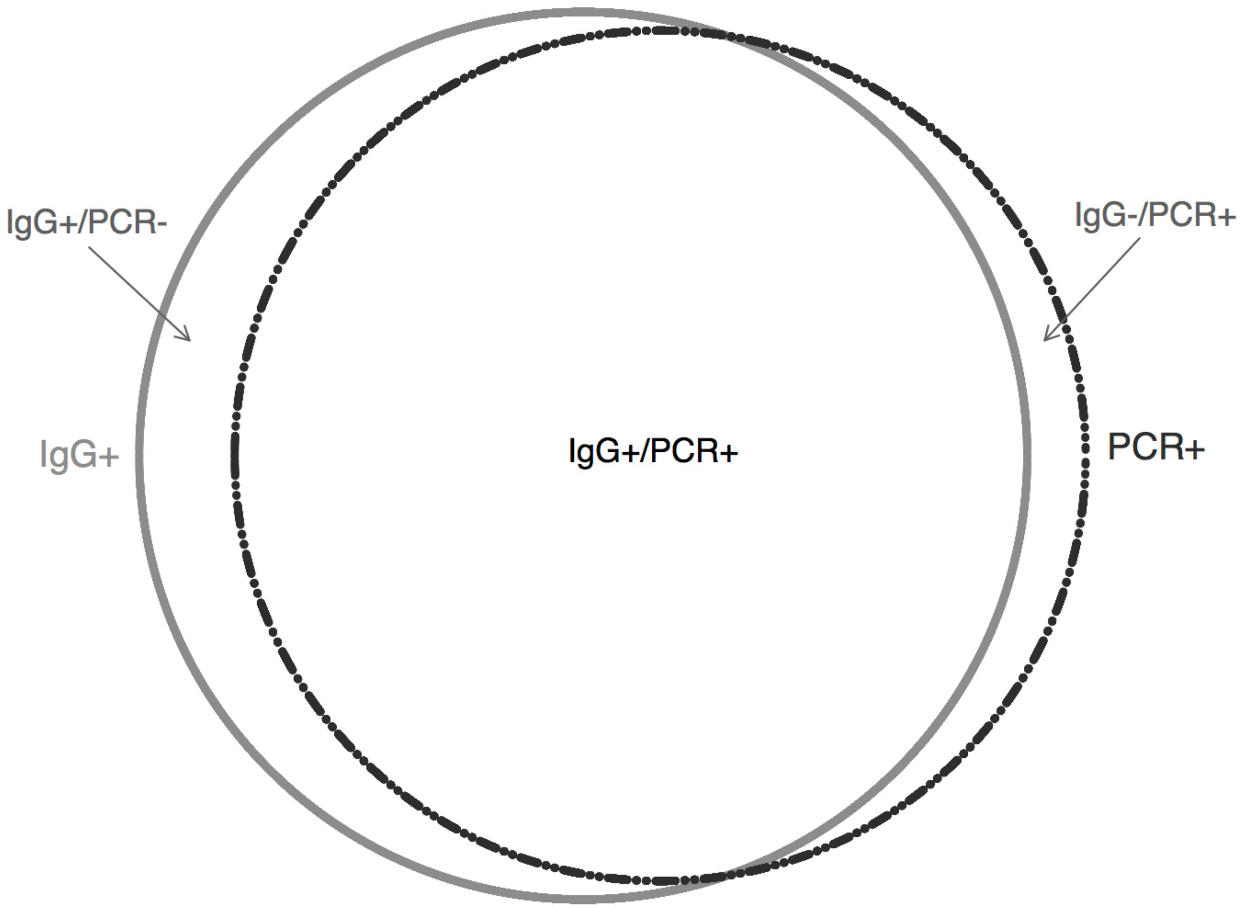
